# The relationship between cognitive function and sleep duration: a Mendelian randomisation study

**DOI:** 10.1101/2020.09.08.20190611

**Authors:** Antoine Salzmann, Nish Chaturvedi, Victoria Garfield

## Abstract

**Importance:** Sleep duration is associated with cognitive function, with Mendelian randomisation evidence supporting a relationship in this direction. However, whether cognitive function may also precede problematic sleep duration remains unclear.

**Objective:** To assess whether reaction time and visual memory are causally associated with sleep duration.

*Design:* Summary-level Mendelian randomisation design between visual memory (30 SNPs), reaction time (44 SNPs), and self-reported and objective sleep duration Setting: Population-based study.

*Participants:* Individuals from the UK Biobank, who were included in genome-wide association studies for our exposures and outcomes, aged 40-69y at baseline (mean 56y), 54% female and self-reported sleep was 7.2 hours.

**Exposures:** Visual memory, reactiontime

**Main outcomes:** self-reported and objective sleep duration

**Results:** Mendelian randomisation results showed that worse performance on the visual memory task was associated with longer (ß=0.09, 95% CI=0.02;0.17), while slower reaction time was associated with shorter (ß=-0.15, 95% CI=-0.29;-0.01), objective sleep duration. Sensitivity analyses revealed no issues with horizontal pleiotropy (MR-Egger intercept p-value <0.05). No association was observed between either cognitive measure and self-reported sleep duration.

**Conclusions and relevance:** These results suggest a potential causative relationship between reaction time and objective sleep duration, where worse visual memory is associated with longer, and worse reaction time with shorter objective sleep duration. This study furthers our understanding of the relationship between brain health and sleep duration and sheds light on the causal nature of these associations.

**Key points:** *Question:* Is genetically predicted sleep duration (accelerometer-derived and self-reported) associated with cognitive function outcomes?

*Findings:* In this summary-level Mendelian randomisation study, worse visual memory was associated with longer, whereas worse reaction time was associated with shorter, objective sleep duration. No associations were observed between cognitive measures and self-reported sleep duration.

*Meaning:* These findings suggest a causal association between cognition and objective sleep duration measures, expanding our understanding of the relationship between cognition and sleep.

## Introduction

Both short and long sleep duration have been associated with adverse cognitive function, cognitive decline and increased risk of dementia in numerous observational epidemiological studies, but findings are somewhat equivocal due to common problems such as reverse causality (e.g. poor cognition leading to disturbed sleep patterns) and inability to fully account for factors that both influence sleep duration and cognition (e.g. adverse mental health) ^1,2^. Given the high prevalence of sleep problems in the population^3^, as well as the public health importance of cognitive impairment and dementia^4^, it is important to understand the direction of these relationships and whether they are causal in nature.

Designs such as Mendelian randomisation (MR) help to overcome some of the limitations of observational studies^6^, due to the fact that genes are randomly allocated at conception and, by nature, phenotypes cannot affect individuals’ genotypes^6^. One previous MR study investigated the causal relationship between self-reported sleep duration and cognitive function, decline, all-cause dementia and Alzheimer’s disease (AD)^7^. Findings suggested that sleep duration was causally associated with cognitive function, but not with cognitive decline, dementia or AD. Specifically, the authors found a U-shaped relationship, such that both short and long sleep duration were associated with worse cognitive function. However, questions remain around bidirectionality. There is growing evidence, from observational studies, that poorer cognitive function may play a role in problematic sleep^8^; thus, providing support for the hypothesis that disordered sleep may be both cause and consequence of adverse cognitive function and dementia^9^. Furthermore, MR studies have shown a unidirectional relationship between increased genetic liability to AD and shorter sleep duration^10,11^, yet no MR studies have investigated the association between cognitive function per se and duration of sleep.

In the present study, we used MR to investigate whether cognitive function is causally related to sleep duration. Genetically predicted cognitive function was measured using visual memory and reaction time tests whilst sleep duration was measured using both self-reported and accelerometer-derived (objective) sleep duration measures. The inclusion of objective sleep measures may increase precision and reduce risk of bias and misclassification as compared to self-reported measures^10,12^. We leveraged data from large-scale summary-level genome-wide association studies (GWAS) of cognitive ability and sleep duration to answer this question.

## Methods

### Study design

We used a pseudo two-sample MR design^13^ to estimate the causal association between cognitive function and sleep duration, using summary data from previously published GWA studies (detailed below). Maximum Ns for our analyses were 446,118 (self-reported sleep duration) and 85,499 (objective sleep duration) UK Biobank (UKB) participants. Our study had sample overlap with the cognitive ability GWA studies which contained UKB individuals. However, for the objective sleep duration outcome subsample, the sample overlap was substantially less (17%) than for self-reported sleep duration.

### Summary statistics and selection of genetic instruments for reaction time and visual memory (exposures)

GWAS summary statistics for 44 reaction time (milliseconds) and 30 visual memory SNPs were downloaded from: http://www.ccace.ed.ac.uk/node/335 and http://www.nealelab.is/uk-biobank, respectively. The reaction time GWAS consisted of 330,069 UKB individuals who completed this test at baseline (time taken to correctly identify matching pairs of cards in a game of ‘Snap’ with higher values indicating worse reaction time) and passed genetic quality control. The visual memory GWAS was performed by the Neale Lab in 361,194 UKB European ancestry participants. We downloaded summary statistics for the raw (untransformed) visual memory test, which corresponded to the number of incorrect matches (higher=worse visual memory) when identifying matches from six pairs of cards after memorising their positions on the screen. We used linkage disequilibrium (LD) clumping in PLINK 1.9 to ensure that SNPs were independent for both reaction time and visual memory (r^2^=0.1, 250kb).

The reaction time and visual memory genetic instruments were both of adequate strength, with F-statistics of 24 (R^2^=0.3%)^14^ and 36.4 (estimated using F=(β^2^/SE^2^)), respectively. For all genetic variants, where the effect/reference alleles were mismatched between the GWASs for our exposures and our outcome, we multiplied the beta coefficient in the sleep duration (outcome) summary statistics by -1 to ensure correct alignment of effect alleles. Details of the reaction time and visual memory SNPs are presented in Supplementary Tables 1 and 2.

### Summary statistics of self-reported and objective sleep duration (outcomes)

GWAS summary statistics were downloaded for both self-reported and objective sleep duration^15^ from the Sleep Disorder Knowledge Portal (http://sleepdisordergenetics.org) and extracted betas and standard errors for the reaction time and visual memory SNPs. Briefly, the self-reported and the objective sleep duration GWA studies comprised 446,118 and 85,499 UK Biobank participants of white European ancestry, respectively. Self-reported sleep duration was based on a question asked at baseline in UKB, specifically ‘About how many hours sleep do you get in every 24h? (please include naps), with responses in hour increments’. Objective sleep duration was derived from accelerometery data collected in a UKB subsample over a period of up to 7 days (median duration 6.9 days). Sleep episodes (defined as periods of at least 5 minutes with no change larger than 5° in the z-axis, which is perpendicular to the accelerometer’s screen and points up) were summed to form a measure of sleep duration^12,15^.

### MR assumptions

MR relies on three core assumptions: i) genetic variants should be robustly associated with the exposure under study (e.g. reaction time and visual memory); ii) genetic variants should be independent of unobserved confounding factors of the relationship under study (e.g. body mass index); iii) genetic variants for the exposure should not directly be associated with the outcome (e.g. reaction time and visual memory SNPs are not associated with sleep duration). Assumption i was met in our analyses, as our genetic variants came from published large-scale GWAS. Assumption ii was tested by uploading our lists of SNPs (for visual memory and reaction time) to PhenoScanner (http://www.phenoscanner.medschl.cam.ac.uk/) and, using p-value thresholds of 0.002 (visual memory = 0.05/30 SNPs) and 0.001 (reaction time = 0.05/44 SNPs) to account for multiple testing, searched for and downloaded associations between the SNPs and the following confounders: BMI, years of education, smoking status, alcohol consumption, systolic blood pressure, diabetes, cardiovascular disease (stroke/coronary heart disease) and hypertension. If a SNP was associated with any of these confounders, then we performed leave-one-out analyses to assess the impact of such SNPs on our MR results.

### Statistical analyses

Analyses were performed in R (version 3.5.2) using the MendelianRandomization package. We performed inverse-variance weighted (IVW) MR as our main model for estimation of causality. This method calculates the effect of the exposure (e.g. reaction time) on the outcome of interest (e.g. self-reported sleep duration) by averaging the ratio of SNP-outcome (*SNP → Y*) to SNP-exposure (*SNP → X*) relationship, calculated using principles identical to a fixed-effects meta-analysis^16^. We also implemented standard MR sensitivity analyses for horizontal pleiotropy, such as MR-Egger regression (provides an intercept term which reveals the extent of unbalanced horizontal pleiotropy)^17^ and the weighted median estimator (WME – yields more robust estimates when up to 50% of the genetic variants are invalid)^18^.

## Results

Participants included in the GWA studies for which we exploited summary statistics were from UKB and thus, aged between 40-69 years at recruitment.

**Table 1.**
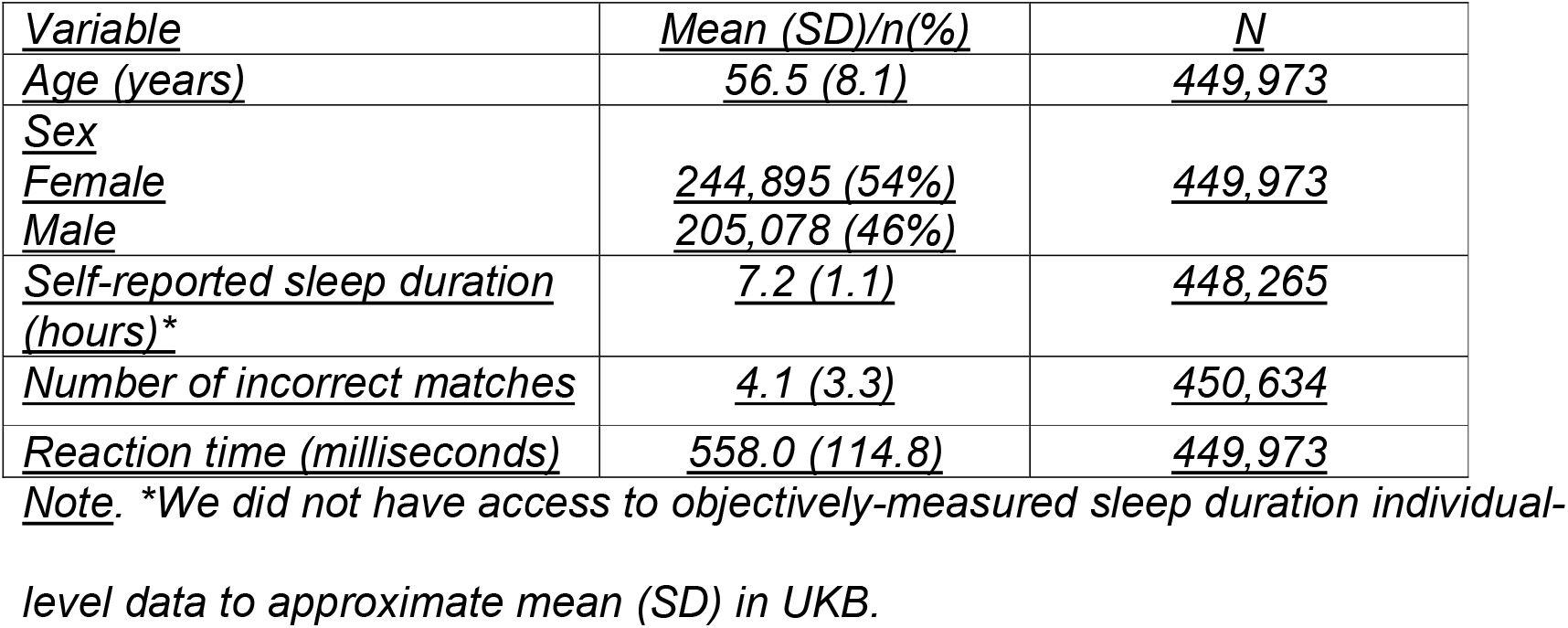
Sample characteristics.

| <u>Variable</u> | <u>Mean (SD)/n(%)</u> | <u>N</u> |
| --- | --- | --- |
| <u>Age (years)</u> | <u>56.5 (8.1)</u> | <u>449,973</u> |
| <u>Sex</u> |  |  |
| <u>Female</u> | <u>244,895 (54%)</u> | <u>449,973</u> |
| <u>Male</u> | <u>205,078 (46%)</u> |  |
| <u>Self-reported sleep duration (hours)*</u> | <u>7.2 (1.1)</u> | <u>448,265</u> |
| <u>Number of incorrect matches</u> | <u>4.1 (3.3)</u> | <u>450,634</u> |
| <u>Reaction time (milliseconds)</u> | <u>558.0 (114.8)</u> | <u>449,973</u> |

Note. \*We did not have access to objectively-measured sleep duration individual-level data to approximate mean (SD) in UKB.

### Visual memory and sleep duration

Results from IVW showed evidence of an association between worse visual memory and longer objective sleep duration (ß=0.09, 95% CI=0.02;0.17). However, this association was weaker in MR-Egger (ß=0.01, 95% CI=-0.50;0.51) and remained identical to the IVW when we ran WME analyses (ß=0.08, 95% CI=-0.01;0.17) (Figure 1). There was however no evidence of an association between visual memory and self-reported sleep duration using IVW (ß=-0.04, 95% CI=-0.12;0.03), MR-Egger (ß=0.17, 95% CI=-0.32;0.65) and WME (ß=-0.02, 95% CI=-0.08;0.04) (Figure 1). There was no evidence of horizontal pleiotropy in any of these analyses (MR-Egger intercept p-value>0.05).

**Figure 1:**
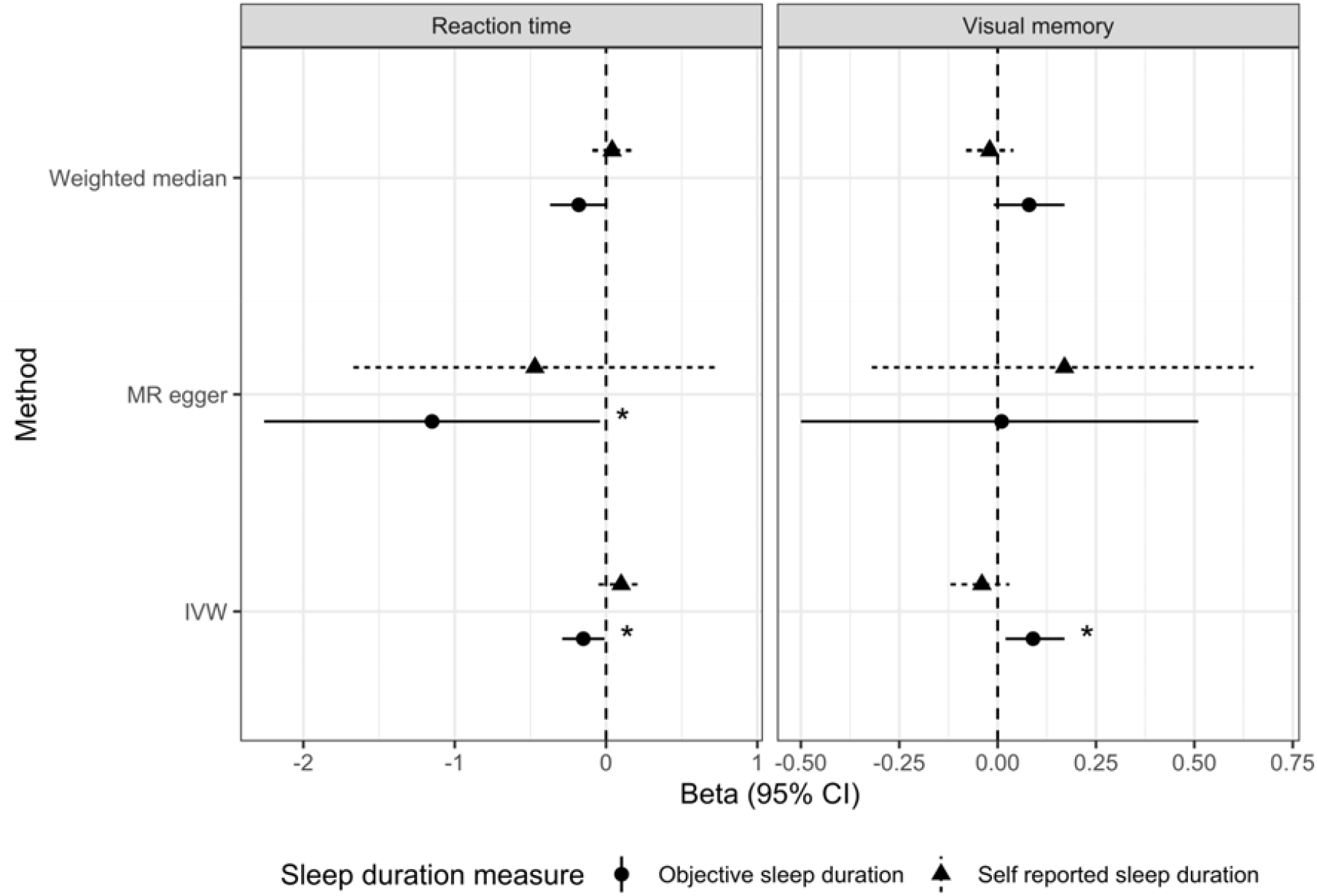
Associations between visual memory and reaction time, and objective and self-reported sleep duration. Abbreviations: Inverse variance weighted (IVW). *Estimate significant at P < 0.05; **significant at P < 0.01.

### Reaction time and sleep duration

MR IVW (ß=-0.15, 95% CI=-0.29;-0.01), Egger (ß=-1.15, 95% CI=-2.26;-0.04) and WME analyses (ß=-0.18, 95% CI=-0.37;0.01) all showed evidence of an association between slower reaction time and shorter objective sleep duration (Figure 1).

However, all MR approaches showed much weaker associations between reaction time and self-reported sleep duration, IVW (ß=0.10, 95% CI=-0.05;0.24), MR-Egger (ß=-0.47, 95% CI=-1.67;0.73) and WME (ß=0.04, 95% CI=-0.09;0.17) (Figure 1). All MR-Egger intercept p-values indicated that there were no concerns with unbalanced horizontal pleiotropy (all p>0.05).

### Additional sensitivity analyses

#### Leave-one-out analyses: visual memory and reaction time SNPs associated with confounders

In these analyses, we excluded SNPs for visual memory and reaction time that were strongly associated with known confounders. SNPs excluded for sensitivity analysis are shown in Supplementary Table 3 and 4. We re-ran all the MR analyses (IVW, MR-Egger and WME) each time we excluded a SNP to assess the impact on our results. Exclusion of each SNP for reaction time and visual memory showed no qualitative difference in the results for objective and self-reported sleep duration (Supplementary Table 1 and 2, respectively).

## Discussion

Using complementary MR approaches, we present two key findings. Firstly, we show that worse visual memory is associated with longer objectively-measured sleep duration with around 5 minutes’ longer sleep for every incorrect match on this task. These results were identical across the IVW and WME approaches, weaker in MR-Egger analyses, yet directionally consistent.

Secondly, we show that slower (worse) reaction time is associated with shorter objective sleep duration. These findings were largely consistent across all MR approaches in terms of the magnitude of the effect (slower reaction time conferred around 9 minutes’ shorter sleep duration in the main MR analysis using IVW), but the WME result did not reach conventional levels of statistical significance. However, our MR analyses showed no evidence of associations between either visual memory or reaction time and self-reported sleep duration.

Interestingly, we also observed that each of the cognitive function measures appeared to have an opposite effect on objective sleep duration, both of which appeared to be quite substantial. In line with our findings, MR studies found increased genetic liability to AD to be associated with shorter self-reported and objective sleep duration^10,11^. While slowing reaction time is a hallmark of normal cognitive ageing, it has long been suggested as an early sign of AD^19^ and our study now shows evidence that it has a negative impact on sleep duration in mid-life. Future work investigating the potential mediating role of cognitive traits on the relationship between AD and sleep may help elucidate the nature of this relationship.

Worse performance on the visual memory task, however, conferred 5 minutes’ longer duration of accelerometer-measured sleep, an estimate that remained consistent across IVW and WME MR approaches. Reaction time was associated with 9 minutes’ shorter sleep duration, which was also identical using IVW and sensitivity WME approaches. Visual memory having a potential effect on longer sleep duration is important because more hours of sleep is not always recommended. For example, both long (>8 or 9 hours, depending on the study) and short sleep duration (<6 or 7 hours) have been associated with increased risk of cardiovascular disease and mortality^20^. However, there is much more evidence linking short sleep duration to poorer physical health outcomes, such as type-2 diabetes, hypertension, obesity^21^, as well as mental health outcomes such as depression and anxiety, via its effect on brain function^22^. That we observed associations in different directions for each cognitive measure and objective sleep duration warrants discussion. In the UKB, visual memory and reaction at baseline are only weakly correlated (Pearson’s r=0.14), indicating that these cognitive domains are somewhat distinctive. This is supported by large-scale population-based studies which show that for example, the association between type-2 diabetes and reaction time is smaller in magnitude than the association between hypertension and reaction time. However, for diabetes vs hypertension and visual memory, neither of these risk factors are strongly associated with this cognitive domain after accounting for confounding factors.

We observed that both visual memory and reaction time were strongly associated with objective sleep duration. This is supported by research which suggests that poorer cognitive function may play a role in problematic (objectively-measured) sleep, such that β-amyloid load in the medial prefrontal cortex (mPFC) is associated with non-rapid eye movement slow wave activity (NREM SWA) impairment^8^. The authors used polysomnography (PSG) to measure sleep parameters in adults without cognitive impairment and found that greater burden of β-amyloid was related to diminished NREM SWA. However, this study was cross-sectional, had a small sample (N=26) and was correlational, which precluded causal inference. Depressive symptoms and the serotonergic system may also play a role in mediating the observed relationship between cognition and sleep. Depression has been commonly linked to impaired attention and memory, in addition to sleep dysregulation. Furthermore, observational studies have previously implicated depression as a mediator of the sleep – cognition relationship.

The only previous MR study of sleep duration and cognitive function used the same cognition phenotypes as the present study, albeit as outcomes^23^. Finding that this relationship is bidirectional and likely causal is important from a clinical perspective. Interventions may target sleep duration as a modifiable lifestyle factor in individuals who have problems with either short/long duration of sleep and this could perhaps improve cognitive function. However, directly targeting cognitive function may prove more challenging.

That we show different results for objective vs. self-reported sleep duration warrants discussion. Firstly, the discrepancies in findings for objective vs. self-reported sleep duration may reflect measurement error and the fact that the agreement between these types of measure is reportedly modest (r=0.45), at best^24^. However, this Pearson’s correlation comes from other datasets, as we did not have access to the objective sleep duration data in UKB. Overall, the results for cognitive function and objective duration of sleep may be more accurate. Secondly, the overlap in samples between the objective sleep duration GWAS and the cognitive function GWAS is substantially lower. This is because, although all of these GWASs included UKB, the objective sleep duration GWAS only had data available on 17% of the sample, as the majority of UKB participants have not undergone actigraphy whereas the self-reported sleep duration GWAS included 90% of the UKB sample. As weak instrument bias is proportional to instrument strength and sample overlap, estimates from analyses utilising the self-reported outcome may be more prone to bias in the direction of the observationally confounded estimate.

Limitations of the present study include the partial overlap in samples in the GWAS summary statistics used. In addition, whilst previous studies have found U-shaped associations (sleep duration -> cognition), the use of summary-level data prevented the investigation of non-linear relationships in our study^23^. Important strengths are 15 the use of objective sleep duration as an outcome; application of a range of MR approaches, alongside additional sensitivity analyses and assumption checks which confirmed the robustness of our results; exploitation of large-scale genomic summary statistics to help disentangle the direction of effect between sleep duration and cognitive function.

In conclusion, using MR, we show that worse reaction time (slower) relates to shorter objective sleep duration, while worse visual memory confers longer objective sleep duration. These results highlight the importance of using objective sleep measurements when trying to understand their relationships with cognitive function and neurological diseases. Furthermore, we observed disctict directions of effect when instrumenting reaction time and visual memory encouraging future work to instrument multiple individual cognitive domains. Overall, our findings contribute to understanding how problematic sleep may be a consequence and not only a cause of, poorer cognitive function.

## Data Availability

The study uses publicly available data from published Genome Wide Associations Studies.

## Acknowledgements

VG is funded by a joint Diabetes UK/British Heart Foundation grant (15/0005250). AS and NC are supported by the UK Medical Research Council (MC_ST_LHA_2019, MC_UU_0019/2).

## Author contributions

VG conceived the study idea and design. AS and VG performed statistical analyses. VG and AS wrote the manuscript; NC provided important intellectual input. All authors approved the final manuscript.

